# Somatic *TET2* Mutations are Associated with Giant Cell Arteritis

**DOI:** 10.1101/2023.07.26.23292945

**Authors:** Michelle L. Robinette, Lachelle D. Weeks, Ryan J. Kramer, Mridul Agrawal, Christopher J. Gibson, Zhi Yu, Aswin Sekar, Arnav Mehta, Abhishek Niroula, Jared T. Brown, Gregory C. McDermott, Edith R. Reshef, Jonathan E Lu, Victor D. Liou, Carolina A. Chiou, Pradeep Natarajan, Suzanne K. Freitag, Deepak A. Rao, Benjamin L. Ebert

## Abstract

**Objective:** Giant cell arteritis (GCA) is an age-related vasculitis. Prior studies have identified an association between GCA and hematologic malignancies (HM). How the presence of somatic mutations which drive development of HM, or clonal hematopoiesis (CH), may influence clinical outcomes in GCA is not well understood.

**Methods:** To examine an association between CH and GCA, we analyzed sequenced exomes of 470960 UK Biobank participants for the presence of CH and used multivariable Cox regression. To examine the clinical phenotype of GCA in patients with and without somatic mutations across the spectrum of CH to HM, we performed targeted sequencing of blood samples and electronic health record review on 114 patients with GCA seen at our institution. We then examined associations between specific clonal mutations and GCA disease manifestations.

**Results:** UKB participants with CH had a 1.48-fold increased risk of incident GCA compared to UKB participants without CH. GCA risk was highest among individuals with cytopenia (HR 2.98, p =0.00178) and with *TET2* mutation (HR 2.02, p =0.00116). Mutations were detected in 27.2% of our institutional GCA cohort, 3 of whom had HM at GCA diagnosis. *TET2* mutations were associated with vision loss in patients with GCA (OR 4.33, p = 0.047).

**Conclusions:** CH increases risk for development of GCA in a genotype-specific fashion, with greatest risk being conferred by the presence of mutations in *TET2*. Somatic *TET2* mutations likewise increase the risk of GCA-associated vision loss. Integration of somatic genetic testing in GCA diagnostics may be warranted in the future.

## Introduction

Giant cell arteritis (GCA) is a systemic granulomatous vasculitis that affects people over age 50 [1]. Due to the risk of permanent vision loss caused by ischemic damage to the ophthalmic vasculature, GCA is a medical emergency that requires rapid recognition and treatment with high dose steroids. IL-6R blockade with tocilizumab (TCZ) is currently the only FDA-approved steroid-sparing therapy [1].

GCA initiating events are largely unknown, but myeloid cell activation may play a central role based on translational, pathologic, and epidemiologic studies [1]. In patients with GCA, circulating monocytes express elevated levels of *IL6* and *IL1B* and are the primary hematopoietic source of systemically elevated IL-6, and neutrophil expansion is the most common cellular abnormality in untreated disease [1, 2]. Myeloid neoplasms (MN) are also associated with GCA. In retrospective studies of patients with inflammatory features of myelodysplastic syndrome (MDS) and chronic myelomonocytic leukemia (CMML), vasculitis is the most common diagnosis, and GCA is the most frequent vasculitis subtype observed [3, 4].

Clonal hematopoiesis (CH) describes the age-related expansion of a clonal population of hematopoietic stem cells and their progeny that are detectable using next generation sequencing, typically after age 50. CH is often caused by somatic mutations in MN driver genes. When the variant allele fraction (VAF) is ≥ 0.02 without evidence of hematologic malignancy (HM), the terms clonal hematopoiesis of indeterminant potential (CHIP) and clonal cytopenia of uncertain significance (CCUS) are used to refer to patients without and with unexplained cytopenia, respectively [5]. CHIP and CCUS have variable risk of evolution to overt myeloid malignancies, including MDS, CMML, and acute myeloid leukemia (AML), with risk determined by hematologic and molecular features [6].

Along with increased risk of MN, patients with CHIP/CCUS have elevated mortality from ischemic cardiovascular disease and develop other age-associated inflammatory diseases [7]. While risk of MN is known to be genotype-dependent, genotype-specific risks for inflammatory diseases are emerging [7]. Mutations in the epigenetic modifier *TET2* may confer a particular risk for inflammatory sequalae, as suggested by retrospective human data and preclinical mouse models which have linked *Tet2*-mutated CHIP/CCUS to ischemic cardiovascular disease, gout, chronic liver disease, and several other inflammatory diseases via myeloid activation [7-11].

Due to the shared associations of CH and GCA with older age and MN as well as mutual evidence of myeloid activation, we hypothesized that CHIP and CCUS would be associated with incident GCA. We further sought to investigate whether the presence of somatic mutations, especially in *TET2*, would influence adverse outcomes in patients with GCA, including incident vision loss and HM. To test this hypothesis, we analyzed data from the UK Biobank, a large population dataset, and a separate GCA patient cohort with deep clinical annotation from our own institution.

## Methods

### UK Biobank cohort

UK Biobank (UKB) data were extracted under application 50834 from a cohort of 502,490 healthy adults 40-70 years of age recruited between 2006-2010. Whole exome sequencing data was analyzed for CHIP-defining somatic mutations as previously described [11]. Individuals with low abundance clones (VAF <0.02), missing laboratory values, and MN diagnosed before or up to 6 months following study enrollment were excluded from this analysis. After exclusions, 470,960 individuals were eligible for study inclusion, including 29,835 with CHIP/CCUS and 441,125 without CHIP/CCUS (Supplemental Figure 1). Incident GCA was identified using linked electronic medical record (EMR) data and international classification of diseases (ICD)-10 codes M31.5 and M31.6.

### Massachusetts General Brigham (MGB) GCA cohort

The MGB cohort of GCA includes 114 individuals that received longitudinal care at MGB; 99% of cases met 2022 ACR/EULAR GCA Classification Criteria. The cohort was identified through ICD-10 code M31.5 and M31.6 search of the MGB Biobank, which has banked DNA samples from >80000 adults across the Mass General Brigham healthcare system in Boston, Massachusetts. EMR abstraction of 387 individuals revealed 98 had positive temporal artery biopsy (TAB), imaging diagnosis of GCA, or clinical diagnosis meeting 1990 ACR GCA Classification criteria diagnosed by a clinical rheumatologist within 6 years of blood sample collection. The second portion of the cohort (n = 16) was prospectively recruited at time of temporal artery biopsy from Massachusetts Eye and Ear and Brigham and Women’s Hospital. EMR abstraction, blinded to genetic data, was performed to annotate patient demographics, GCA outcomes, medication use, blood counts, referral to hematology, and development of HM. Targeted next generation sequencing, alignment, and genetic variant calling was performed as previously described [12] using the Illumina platform (California, USA) and libraries generated with two custom hybrid capture probe sets from Twist Biosciences (California, USA). All subjects provided written informed consent to participate in biobanking. The IRB of Mass General Brigham gave ethical approval for biobanking and EMR abstraction protocols. For extended details on cohort identification, EMR abstraction, and sequencing methods, see Supplemental Methods.

### Cell sorting and DNA extraction

Cryopreserved peripheral blood mononuclear cell (PBMC) were available from four prospectively recruited individuals with somatic mutations and were sorted into cell fractions as previously described [13]. For gating strategy and reagents, see Supplemental Methods. The QIAamp DNA Blood Mini kit (Qiagen, 51104) and the Zymo DNA Microprep Kit (D3021) were used for DNA extraction from bulk and sorted samples, respectively.

### Statistical Analyses

Statistical analyses and figure preparation were performed using R (R Foundation for Statistical Computing) and GraphPad Prism (GraphPad Software). Wilcoxon rank sum test and Fisher’s exact tests were used to evaluate continuous and categorical data, respectively. Cox proportional hazards regression analyses were performed to determine hazard ratios (HR) for incident GCA and 95% confidence intervals. Wilcoxon matched-pair signed rank test was used for paired values in mutation segregation. Benjamini & Hochberg correction was applied to multiple comparisons and adjusted p values (p-adj) are noted. Odds ratios (OR) for vision loss were assessed using Fisher’s exact test and the Baptista-Pike method for confidence intervals. The threshold for statistical significance was p/p-adj ≤ 0.05.

For detailed methods, see Supplemental Methods.

## Results

### CHIP/CCUS is associated with incident GCA in the UKB

A total of 779 out of 470,960 (0.165%) individuals in the UKB had incident GCA. Of these, 82 individuals with incident GCA had CHIP/CCUS and 697 did not (Supplemental Figure 1). Multivariable Cox proportional hazards models adjusted for age, sex, and smoking history were performed to compare the risk of incident GCA in individuals with and without CHIP/CCUS. The hazard ratio for incident GCA was 1.47 (95% confidence interval 1.17 -1.856, p = 0.000927) in CHIP/CCUS relative to controls. The risk of incident GCA was 2.98-fold higher in CCUS (p=0.000178) and 1.68-fold higher in CHIP (p = 0.015) relative to unmutated participants. Of the three most common CHIP/CCUS genotypes (*DNMT3A, TET2*, and *ASXL1*), *TET2* was the only genotype independently associated with increased risk for incident GCA [HR 2.02 (1.32 - 3.10), p = 0.00116] (Figure 1A).

**Figure 1.**
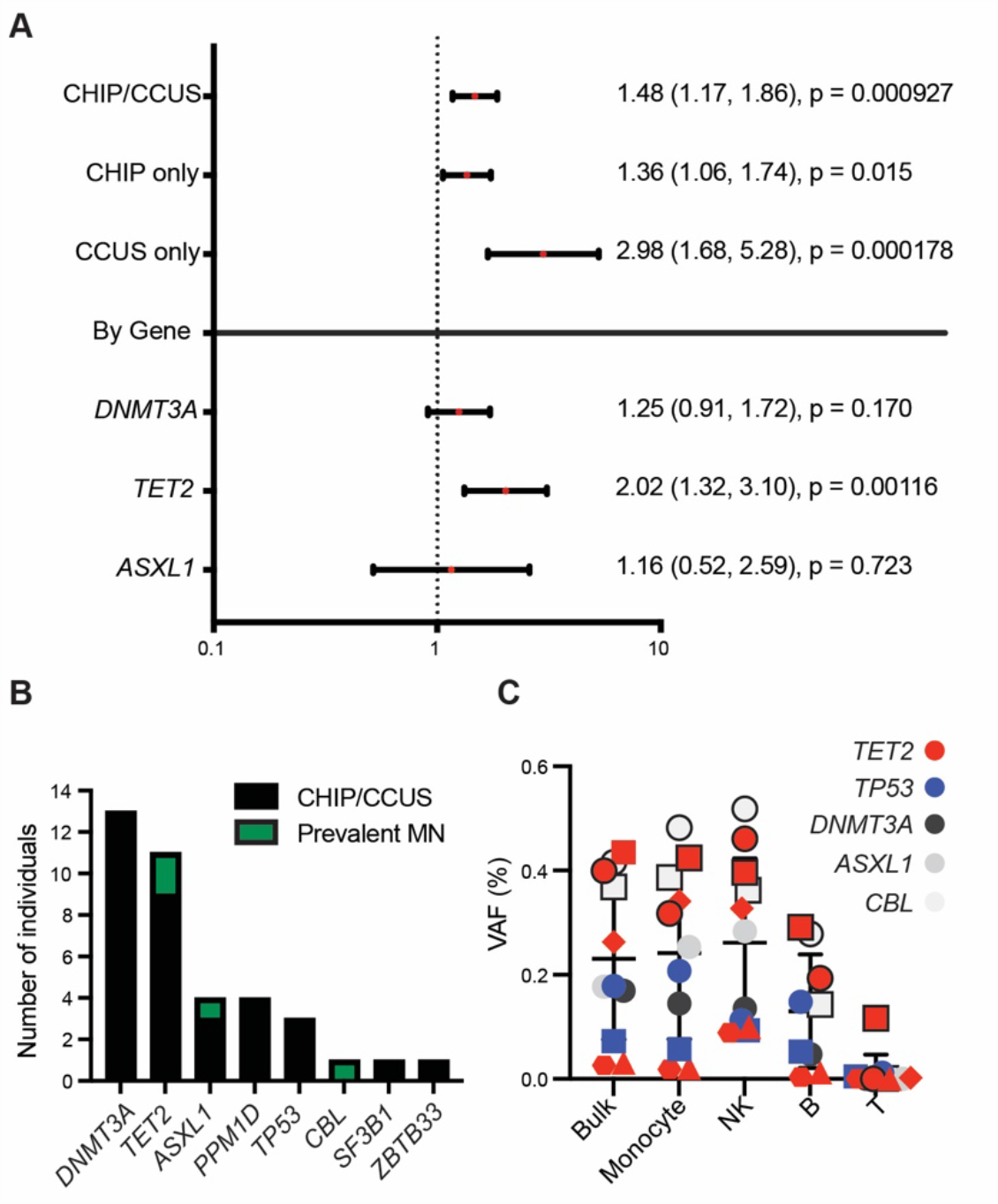
Genetic profile of CH in GCA cohorts. **A**) Multivariable Cox proportional hazards model showing hazard ratios (HR) for incident GCA among individuals from UKB with CHIP/CCUS relative to no CHIP/CCUS; CHIP or CCUS relative to no CHIP/CCUS; and *DNMT3A, TET2*, or *ASXL1*. In all cases, multivariable models were performed with no CHIP/CCUS as the reference group. All models were adjusted for sex, age, and smoking status (any smoking history). Forrest plot displays main effects (HR) with 95% confidence intervals. Numerical values for HR (95% CI) with associated p-values are provided to the right of each graph. For all statistical tests, significance threshold is set at p <0.05. **B)** Stacked bar graph showing number of individuals with mutations ≥0.02 on a per gene basis, where black represents CHIP/CCUS and red prevalent MN Characteristics of CH in GCA from individuals from MGB. **C)** All prospectively recruited individuals with VAF ≥ 0.02 detected on bulk sequencing either had paired PBMC banked from the same whole blood used for initial sequencing (n = 3) or 1 month later (n = 1). One B cell fraction failed QC. Data are shown per mutation, where colors represent the gene mutated and shapes represent distinct mutations per gene. Black outlined blue and gray shapes represent mutations from prevalent MN. Monocyte versus NK cell VAF: CH + MN p-adj = 0.2783, CH only p-adj = 0.5781. Monocyte versus B cell VAF: CH + MN p-adj = 0.0059; CH only p-adj = 0.0937. Monocyte versus T cell VAF: CH + MN p-adj = 0.0030, CH only p-adj = 0.04680.

## CHIP, CCUS, and MN are prevalent in the MGB GCA cohort

We performed a genetic analysis of our cohort of 114 MGB patients with GCA, sequencing genes that are recurrently mutated in MN (Table 1). We detected these mutations in 31 individuals (27.2% of total cohort), including 28 (90.3%) who had CHIP or CCUS and 3 individuals (9.7%) with prevalent MN at GCA diagnosis (Supplemental Table 1). *DNMT3A, TET2* and *ASXL1* mutations were the most common in the MGB cohort (Figure 1B). In four individuals with available PBMC samples (n = 3 CH, n = 1 MN), we flow sorted cells from different hematopoietic lineages and examined the VAF of mutations in each lineage.

**Table 1:** Description of the Confirmed MGB GCA cohort. PMR = Polymyalgia Rheumatica. CHIP = Clonal Hematopoiesis of Indeterminate Potential. CCUS = Clonal Cytopenia of Uncertain Significance. MN = Myeloid Neoplasia.

|  | Total | No CHIP/CCUS | CHIP/CCUS | Prevalent MN |
| --- | --- | --- | --- | --- |
| n | 114 | 83 | 28 | 3 |
| Age at GCA diagnosis<br>median [IQR] | 73.2<br>[65.8, 78.0] | 73.0<br>[64.0, 77.5] | 75.5<br>[70.6, 79.2] | 82.9<br>[75.1, 84.2] |
| Age at sequencing<br>median [IQR] | 74.37<br>[67.5, 80.3] | 74.2<br>[65.8, 78.5] | 76.7<br>[70.4, 81.7] | 84<br>[76.8, 84.8] |
| Years of follow up<br>median [IQR] | 5.75<br>[3.10-8.33] | 5.7<br>[3.1, 8.4] | 6.0<br>[3.17, 7.75] | 3.08<br>[1.67-3.33] |
| Diagnosis Type (n, %) |  |  |  |  |
| Biopsy | 65 (57%) | 45 (54.2%) | 17 (60.7%) | 3 (100%) |
| Imaging | 23 (20.2%) | 17 (20.5%) | 6 (21.4%) | 0 (0%) |
| Clinical | 26 (22.8%) | 21 (25.3%) | 5 (17.9%) | 0 (0%) |
| Male sex (n, %) | 31 (27.2%) | 18 (21.7%) | 11 (39.3%) | 2 (66.7%) |
| Ever smoker (n, %) | 50 (43.9%) | 37 (44.6%) | 12 (42.9) | 1 (33.3%) |
| PMR (n, %) | 63 (55.3%) | 47 (56.6%) | 15 (53.6%) | 1 (33.3%) |
| Vision loss (n, %) | 16 (14.0%) | 10 (12%) | 5 (17.9%) | 1 (33.3%) |

Mutations were detected in all cases in monocytes. Compared to monocytes, mutations were detected at equivalent VAF in NK cells, lower VAF in B cells, and were rarely observed in T cells (Figure 1C).

## Genotype-Specific Outcomes and GCA

We focused on vision loss as it is a severe, binary outcome in patients with GCA. We observed 19 vision loss events in 16 patients (Supplemental Figure 2A). We observed 5 cases of vision loss among individuals with CHIP/CCUS and 1 case of vision loss in prevalent MN (Figure 2A, Supplemental Figure 2B). Of these 6 cases with vision loss and somatic mutations, 4 (66.6%) were in biopsy-proven GCA patients with *TET2* mutations and none had isolated *DNMT3A* mutations (Figure 2A, Supplemental Table 2). Evaluated by genotype, *TET2* mutations were significantly associated with vision loss (CH + MN OR 4.33, 95% CI 1.25-17.8 p = 0.047; CH only OR 3.18, 95% CI 0.7971-12 p = 0.134, Figure 2B). CRP was significantly lower in individuals who had somatic mutations and vision loss (CH+ MN median CRP 12.15 vs 122.0 p = 0.0120; CH only median CRP 13.9 vs 122.0, p = 0.0190, Figure 2C), while ESR was not different (Supplemental Figure 2B). Vision loss was commonly the event precipitating steroid initiation for suspected GCA (Supplemental Table 2, Supplemental Figure 2C). We observed that individuals with vision loss, before starting corticosteroids, had a lower lymphocyte relative and absolute counts (relative lymphocyte count median 9.55% vs 18.15%, p = 0.0042; absolute lymphocyte count median 1.03 vs 1.56 p = 0.0023; Figure 2D). The lower lymphocyte percentage among GCA patients with vision loss was explained by a higher proportion of circulating myeloid cells, of which only the percentage of monocytes was significantly different (Supplemental Figure 2D). Individuals with CHIP/CCUS and vision loss had larger clone sizes, measured by maximum VAF, compared to those without vision loss (median max VAF 0.18 vs 0.0634 p = 0.0453). We observed 2 cases of incident HM, both in patients with *TP53* mutations.

**Figure 2:**
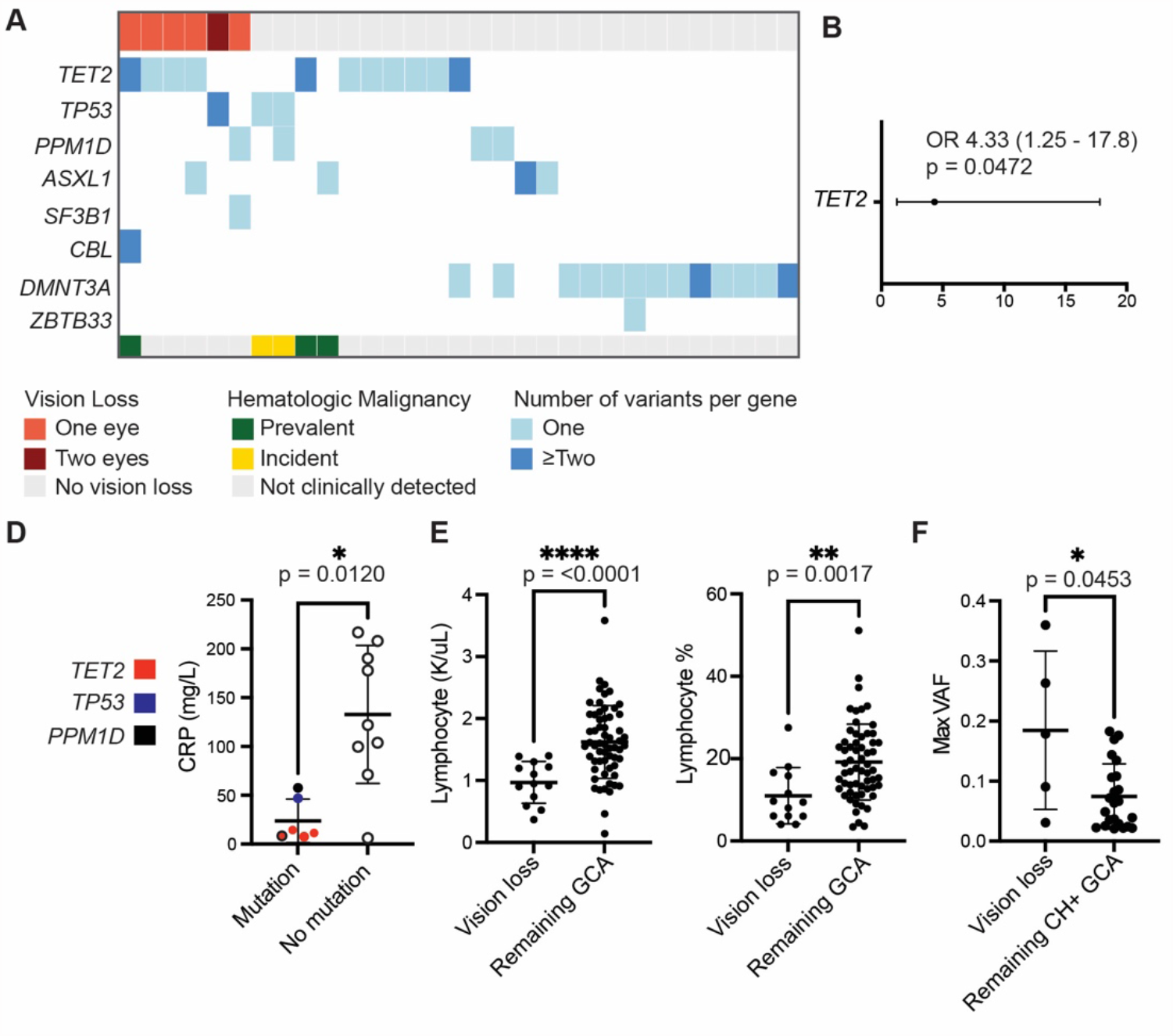
Clinical outcomes in GCA patients with CH. **A)** Co-occurrence matrix of severe outcomes in GCA clustered by outcome and genotype, where individuals are represented by one column. B) Forrest plot demonstrating odds ratio for vision loss with *TET2* mutation across the MGB cohort. **C)** Pre-steroid CRP in GCA patients with vision loss with somatic mutation versus no mutation. **D)** Pre-steroid lymphocyte percentage and absolute lymphocytes counts in GCA patients with vision loss compared to those without vision loss, excluding prevalent MN. **E)** Maximum VAF of individuals with CHIP/CCUS and vision loss, compared to those without vision loss.

Cytopenias, especially anemia, were common at diagnosis and at last follow up, regardless of somatic mutation status (Supplemental Table 3-4). A minority of GCA patients were referred to hematology for cytopenia evaluation, and among those, an even smaller proportion underwent diagnostic evaluation for HM (Supplemental Table 5).

## Discussion

Here, we demonstrate an association between CHIP and incident GCA. More specifically, we find that GCA is more common in *TET2* mutated CHIP/CCUS compared to other genotypes, and that *TET2* somatic mutations are associated with GCA-related vision loss. Analysis of our institutional MGB cohort, though uncontrolled, is consistent with our findings in the UKB. Future prospective investigation of GCA patients with CHIP/CCUS or MN with age and sex-matched controls will allow for a more complete understanding of how mutations in myeloid driver genes influence clinical trajectories and adverse outcomes in this population.

GCA is predominantly a myeloid and CD4^+^ T cell mediated disease. In cases where we examined the presence of somatic mutations in specific hematopoietic lineages, mutations were always present in monocytes but rarely detectable in T cells [7, 13], suggesting that the contribution of somatic mutation to GCA may be mediated by myeloid cells. In other model systems, Tet2-deficient monocytes and macrophages display enhanced response to inflammatory stimuli, especially enriched for increased IL-1B and prolonged IL-6 production [7-9, 11, 14, 15], cytokines well recognized to be elevated in GCA monocytes [2]. Indeed, recent models have emphasized the central role of myeloid cells in the initiation of vascular injury in GCA, based on observations of circulating myeloid activation in GCA patients and experimental evidence that T cells require myeloid cells to enter blood vessels [1].

Features previously associated with increased risk of HM, namely CCUS and larger clone size, were also associated with increased development of GCA and vision loss. Despite these associations, individuals from our institutional MGB cohort with GCA were not frequently evaluated for HM. We have recently shown that the risk of incident MN and all-cause mortality in CHIP/CCUS can be predicted with age, hematologic indices including MCV, RDW, and presence of cytopenia, and sequencing data regarding number, size, and type of genetic mutation [6]. This prognostication may be useful should genetic testing become more commonly used in GCA or other forms of vasculitis in the future. Moreover, as patients with CH are increasingly followed in designated clinics, clinicians should consider GCA in patients with *TET2* mutations and headache even in the absence of substantially elevated inflammatory markers.

## Data Availability

All data produced in the present study are available upon reasonable request to the authors

## Author Contributions

M.L.R, L.D.W, M.A., and B.L.E designed and initiated the project. M.L.R., M.A., A.M., and G.C.M. performed chart review. L.D.W. and Z.Y. performed biobank analyses. C.J.G, A.S., A.N. and P.N. generated somatic mutation calls for UK Biobank and Mass General Brigham Biobank studies. M.L.R, L.D.W, R.J.K., and J.B. analyzed data. M.L.R, E.R.R., J.E.L., V.D.L, C.A.C., S.K.F., and D.A.R. collected samples. M.L.R., L.D.W., and B.L.E. drafted the manuscript; all authors made substantial contributions to data analysis and/or interpretation, drafted and/or revised the manuscript, and approved the final version for publication.

## Acknowledgments

This work was supported by the National Institute of Arthritis and Musculoskeletal and Skin Diseases (T32 AR055855) (M.L.R, G.C.M.); ASH/RWJF AMFDP and Edward P. Evans Foundation for MDS (L.D.W); the Sarnoff Cardiovascular Research Foundation (R.J.K); the Deutsche Forschungsgemeinschaft (DFG, AG252/1-1) (M.A.); Knut and Alice Wallenberg Foundation (no. KAW2017.0436)(A.N.); National Institutes of Health (NIH) K07-CA263555 (C.J.G); the National Heart, Lung, and Blood Institute (5T32HL007604-37) (Z.Y.) and (T32HL116324) and Mittelman Fellowship (A.S.); the Doris Duke Charitable Foundation Physician Scientist Fellowship (M.A.); NHLBI R01HL148050 (P.N.); NIH K08-AR072791, P30-AR070253, and Burroughs Wellcome Fund Career Award in Medical Sciences (D.A.R.); NIH (R01HL082945, P01CA108631, P50CA206963, and R35CA253125), the Howard Hughes Medical Institute, and the Adelson Medical Research Foundation. The authors would additionally like to thank Dr. Joseph A. Rizzo and Dr. Bart K. Chwalisz from MEEI Neuro-ophthalmology; Dr. Sara Tedeschi, Ms. Lin Chen, and Ms. Eilish Dillon from BWH Rheumatology; and Dr. Michael Belkin, Dr. Edwin C. Gravereaux, Dr. Mohamad A. Hussain, Dr. Matthew T. Menard, and Dr. Charles K. Ozaki from BWH Vascular Surgery for additional assistance facilitating patient sample collection and sample pre-processing.

