## Supplemental Data for "Somatic *TET2* Mutations are Associated with Giant Cell Arteritis"

Supplemental File

### Supplemental Methods

#### Cohort composition and sequencing methods

***Massachusetts General Brigham Biobank (MGBB)***
All blood DNA samples were sequenced using a 98-gene custom Twist panel as previously described [1]. Samples were identified from MGBB using the following criteria: 1- ICD-10: M31.5 and M31.6, 2- age > 40, 3- Biobank DNA sample available, resulting in 387 individuals. These were screened by an MD for documentation of GCA in chart, positive temporal artery biopsy (TAB), positive PET or ultrasound, and strong clinical suspicion for GCA with elevated ESR, resulting in 176 cases.

In a second round of chart review, a rheumatologist reviewed the screened charts for GCA, resulting in 117 cases. Inclusion required a positive TAB, imaging diagnosis consistent with diagnosis of GCA, or clinical diagnosis meeting 1990 ACR Classification Criteria made by a clinical rheumatologist. All TAB that had pathology available were additionally reviewed for inflammatory infiltrates and were only considered positive if inflammatory cells were seen or active arteritis noted; if absent, GCA was considered a clinical diagnosis.

To account for temporal connection of CH to GCA, blinded to genotype information, samples were only included if collection occurred within 6 years before or after GCA diagnosis, resulting in 98 cases. This number was chosen based on median the time to incident GCA in UKB analysis of 6.7 years.

***Massachusetts General Brigham Prospective Recruitment***
Between August 2020 and August 2022, patients were prospectively and sequentially enrolled in research at the time of temporal artery biopsy at Massachusetts Eye and Ear Infirmary (MEEI) with consent performed by the provider who performed the procedure, resulting in 57 cases. Additional patients were also prospectively collected from Brigham and Women's Hospital (BWH), resulting in 9 cases. There was no selection for cases, but multiple rheumatologists requested TAB and consent was not performed by the vascular surgery provider who performed the procedure, resulting in lower capture. Of the 66 consented patients, 18 patients had positive TAB, 2 from BWH and 16 from MEEI. Blood was then drawn from these patients at clinical follow up at MEEI or BWH, with 2 lost to follow up from MEEI, resulting in 16 cases.

Samples were sequenced using a 13-gene custom Twist panel as previously described [2]. Sorted cell fractions from samples with CH identified on bulk sequencing were also sequenced using this platform.

***Massachusetts General Brigham Hospital GCA cohort***

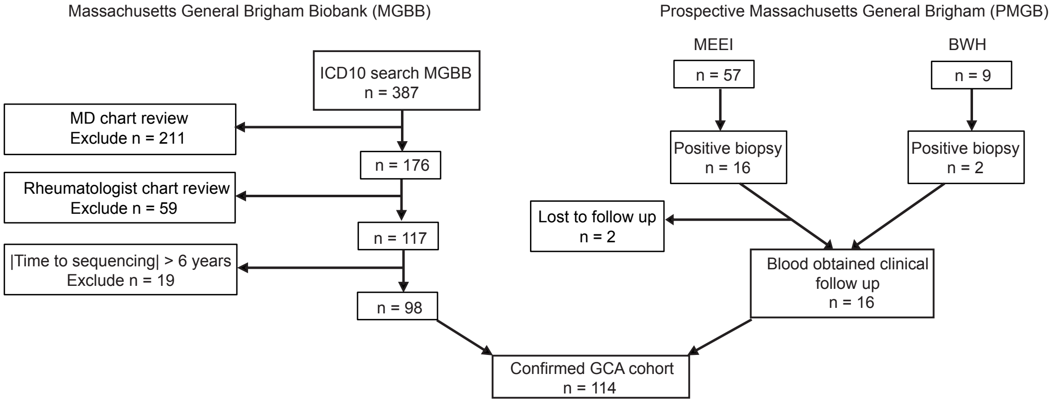


The institutional MGB GCA cohort was a composite cohort that included the above 98 cases from MGBB and 16 prospectively recruited cases. Extended analyses focused only on the shared genes sequenced in both cohorts.

2022 ACR/EULAR Classification Criteria were published after cohort identification [3]. Post-hoc, all cases were reviewed for these criteria. 113 of 114 cases meet classification criteria. The single case that did not meet criteria was a woman in the age bracket 61-65 who had a sum score of 5, with ESR 115 mm/h, CRP 59.6 mg/L, new temporal headache, and diffuse wall thickening on CTa of aorta and branch vessels; FDG PET was not obtained, axillary vessels were not assessed, and TAB was negative. This patient was enrolled in a double-blinded RCT for GCA. This patient was included in subsequent analysis.

**Sequencing Analysis**

Raw sequence data was aligned to hg19 using BWA-mem. Following generation of pileup files using mpileup from samtools version 1.3.1, variant calling was performed with Varscan version 2.3.3 with minimum number of alt reads 5, minimum depth 50, maximum noise of 20. Annotation was performed using Annovar version 2018Apr16, including gnomad_genome and gnomad_exome frequencies. Individual single nucleotide substitutions and small insertions or deletions were evaluated as candidate drivers of clonal hematopoiesis based on gene-specific characteristics, then curated manually and classified as driver mutations based on genetic criteria and literature review, as previously described. [4]

**EMR Abstraction**

Extended electronic medical record abstraction was performed blinded to any genetic data. Demographic data including smoking use and sex were documented. Diagnosis of GCA was considered the date at which point high dose steroids were initiated empirically for suspected diagnosis GCA. Diagnosis type (i.e. biopsy, imaging, or clinical) was documented. All rheumatology and ophthalmology notes were reviewed and any diagnosis of PMR or vision loss were documented. For vision loss episodes, date and type of loss was also documented.

Complete blood counts (CBCs) were extracted from composite of all available documentation in Epic including archival medical records, scanned records, Care Everywhere, and BI Boston for all patients, with CBC choice parameters pre-specified after practice review of 7 random cases, which were then re-evaluated with the same methods with the remaining cohort. All CBC and inflammatory marker dates were documented and were mapped by chart review with documentation of dose of steroid and other pertinent medications including conventional DMARDs, biologics, and chemotherapy, if present. CRP values represent a compositive of CRP and high-sensitivity CRP. Because CBC were not available at the time of genetic sequencing for many individuals, CHIP and CCUS were not able to be differentiated in the MGB cohort.

Initial CBC:
The CBC after charted GCA symptom onset as close to GCA diagnosis prior to steroid initiation was chosen. If an antecedent diagnosis of PMR predated GCA diagnosis leading to steroid initiation, the CBC at PMR diagnosis was used if within 6 months of GCA diagnosis. If pre-steroid CBC were not documented in the time where GCA symptoms clearly existed and led to bloodwork evaluation or PMR diagnosis was >6 months before GCA onset, then the first CBC after GCA diagnosis on steroids was documented. However, only CBC with strict cut off 0 steroids and no steroids in past 30 days were included in further analysis. Inflammatory markers of ESR and CRP were documented with the same strategy, but dates were occasionally different between CBC and thus medication use was documented separately and the same cut off applied. For patients with somatic mutation and vision loss, an external reviewer was provided the chart and date of CBC/ inflammatory markers without further information, then independently reviewed the chart, confirming each patient was not on steroids at the time of laboratory evaluation or in the 30 days prior.

Final CBC:
Representative final CBC was selected from chart review without major confounder, such as admission for sepsis, major injury, terminal hospitalization, active chemotherapy, documented or suspected infection. Preference was made for outpatient CBC if representative values were available and not confounded by post-hospital follow-up; but if unavailable, urgent care or ED visits for minor complaints were accepted. If no CBC with differential was found for ≥2 years, a CBC without differential was selected. All cancers and chemotherapy courses were documented. For patients who developed incident malignancy requiring chemotherapy after GCA, last CBC was used prior to chemotherapy initiation. For individuals who developed GCA while taking chemotherapy (n=3), final CBC was selected prior to additional lines of chemotherapy if required.

Referral to hematology and hematologic malignancy diagnoses:
All charts were systematically reviewed for any hematology and oncology referral. If a referral to hematology was ever made, the date of referral, work up obtained, testing performed, and diagnosis was documented.

**Cell sorting**

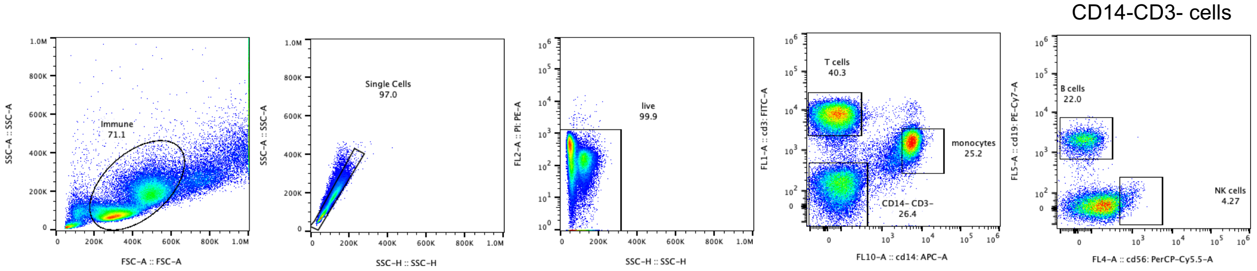


Cryopreserved cells were thawed, washed twice in PBS + 5% FCS, then pre-incubated with BD Pharmingen Fc block (564219) for 10 minutes, to which CD3-FITC BD (555339), BD CD19-PeCy7 (560728), Ebioscience CD14-APC (17-0149-42), and BD CD56-PerCP Cy5.5 (560842) were added for 30 minutes. Cells were washed twice and then sorted.

Sorting gates were established using healthy control sample, with purity gating of sorted fractions all ≥98.8%. Sorting strategy for PBMC from pre-specified gates with a representative patient sample shown above.

1. Miller, P.G., et al., *Contribution of clonal hematopoiesis to adult-onset hemophagocytic lymphohistiocytosis.* Blood, 2020. **136**(26): p. 3051-3055.
2. Miller, P.G., et al., *Clonal hematopoiesis of indeterminate potential and risk of death from COVID-19.* Blood, 2022. **140**(18): p. 1993-1997.
3. Ponte, C., et al., *2022 American College of Rheumatology/EULAR Classification Criteria for Giant Cell Arteritis.* Arthritis Rheumatol, 2022. **74**(12): p. 1881-1889.
4. Gibson, C.J., et al., *Donor Clonal Hematopoiesis and Recipient Outcomes After Transplantation.* J Clin Oncol, 2022. **40**(2): p. 189-201.

Supplemental Tables

Supplemental Table 1: Characteristics of prevalent and incident clinically detected hematologic malignancies

Supplemental Table 2: Characteristics of vision loss occurring greater than 1 week from steroid initiation

Supplemental Table 3: Complete blood counts and cytopenia at GCA diagnosis

Supplemental Table 4: Cytopenia at last follow-up

Supplemental Table 5: Referral to hematology for GCA patients

| Hematologic Malignancy Diagnosis | Time (years) hematologic diagnosis relative to GCA diagnosis | GCA Diagnosis Type | Molecular profile at clinical sequencing | Molecular profile at research sequencing | Time (years) clinical sequencing relative to research sequencing | Treatment of (Prevalent Disease) Prior to Incident Disease |
|---|---|---|---|---|---|---|
| Prevalent Diagnoses compared to GCA | | | | | | |
| MDS<br>&<br>CLPD-NK | -1.5<br><br>-1.5 | Biopsy | *TET2* p.S1612fs 18.0%[#]<br>TET2 p.Q255fs 16.4% [#]<br><br>46,XX[20]<br><br>Source: Blood | *TET2* p.S1612fs 32.1%<br>*TET2* p.Q255fs 25.2%<br>*SRSF2* p.P95H 35.5%<br><br>Source: Blood | -2.6 | (MDS) None<br><br>(CLPD-NK) None |
| MDS | -0.5 | Biopsy | *ASXL1* p.T712fs[&] 18%<br><br>46,XY[20]<br><br>Source: Bone marrow | *ASXL1* p.T711fs 9.1%[&]<br><br>Source: Blood | +1 | (MDS) None |
| CMML | 0 | Biopsy | *CBL* p.R420L 41.6% [#]<br>*CBL* p.R420* 36.9% [#]<br>*PHF6* p.K324E 85.3%<br>*TET2* p.Q1813* 43.5% [#]<br>*TET2* p.K1752Nfs 14 40%[#]<br>*JAK2* p.V617F 1.7%[+]<br><br>45,X,-Y,del(20)(q11.2q13.3)[20]<br>Source: Blood | Bulk sequencing not repeated | -0.08<br><br>(PBMC fractions) | (CMML) None |
| Incident Diagnoses compared to GCA | | | | | | |
| AML | +2.75 | Imaging | 44,XX,-3,t(4;11)(q23;p15),del(5)(q13q31),-16,der(17)t(17;20)(p11.2; p11.2)add(17)(q21),-18,+21,+mar[14]/46,XX[6][o]<br><br>Source: Bone marrow | TP53 p.L330R 10.8%<br><br>Source: Blood | +1.75 | (GCA) Steroids |
| Smoldering Myeloma | +3 | Biopsy | FISH: Monosomy 13<br>46, XX[20]<br><br>Source: Bone marrow | None detected<br><br>Source: Blood | +0.67 | (GCA) Steroids, TCZ |
| DLBCL, germinal center subtype | +4.42 | Biopsy | *TP53* p.R306X 88.3%<br>*IKZF3* p.N160S 35.7%<br>*ATM* p.R2161H 11.7%<br><br>Source: Lymph node | *TP53* p.R306X 14.4%<br>*PPM1D* p.W427X 2.6%<br><br>Source: Blood | +4.9 | (GCA) Steroids |
| Myelodysplastic Syndrome; CLPD-NK = Chronic Lymphoproliferative Disorder of Natural Killer Cells; CMML = Chronic Myelomonocytic Leukemia; AML = Acute Myeloid Leukemia; DLBCL = Diffuse Large B Cell Lymphoma; TCZ = Tocilizumab. [#] The two variants of the same gene are in trans. [+]Variants <2% are less than validated limited of detection. ^Known germline variant. [&]Aligned to hg38 clinically, hg19 for research. [o]Only karyotyping performed. [$]Expired before bone marrow biopsy performed. | | | | | | |

**Supplemental Table 1: Characteristics of prevalent and incident clinically detected hematologic malignancies**

| Time to vision loss from steroid initiation | Time from sequencing to GCA diagnosis (years) and mutations | GCA Diagnosis Type | Vision loss details | GCA treatment course before vision loss |
|---|---|---|---|---|
| -14 days | 0 years<br><br>*CBL* p.R420L 41.6%<br>*CBL* p.R420 36.9%<br>*PHF6* p.K324E 85.3%<br>*TET2* p.Q1813 43.5%<br>*TET2* p.K1752Nfs 14 40%<br>*JAK2* p.V617F 1.7% | Biopsy | AAION<br><br>(Original diagnosis NAION) | No steroids until develops eye pain and photopsia in second eye with jaw claudication |
| +8 days | +0.6 years<br><br>*TP53* p.R282G 17.9%<br>*TP53* p.R273C 7.2% | Biopsy | Second eye AAION<br>(Originally suspected NAION) | Prednisone 100 mg - 2 days<br>Prednisone 60 mg - 6 days |
| +15 days | -6 years<br>*TET2* p.Q918X 36% | Biopsy | PION | Prednisone 50 mg - 15 days |
| +27 days | +2.3 years<br><br>No variant detected | Clinical | AAION<br><br>(Original diagnosis PMR) | Prednisone 40 mg - 4 days<br>Prednisone 20 mg - 12 days<br>Prednisone 10 mg - 2 days<br>Prednisone 20 mg - 9 days |
| +54 days | +1 year<br><br>No variant detected | Biopsy | Second eye AAION | IV Solumedrol - 5 days<br>Prednisone 80 mg - 36 days<br>Prednisone 60 mg - 13 days |
| +231 days | + 0.2 year<br><br>*PPM1D* p.W247X 9.2%<br>*SF3B1* p.E622D 6.6% | Clinical | AAION | Prednisone 80 mg - 2 days<br>IV solumedrol - 3 days<br>Prednisone 80 mg ~2 weeks<br>Prednisone 60 mg ~1 week<br>Prednisone 40 mg -> tapered by 5 mg every 2 weeks -> 20 mg<br>Prednisone 20 mg -> tapered by 2.5 mg every 2 weeks -> 10 mg<br>Prednisone 10 -> taper to 5 mg over two months<br>Prednisone 1 mg (in error) ~1 week<br>Prednisone 40 mg x 2 days |

**Supplemental Table 2: Characteristics of vision loss occurring greater than 1 week from steroid initiation**

| | No CHIP/CCUS | CHIP/CCUS | Prevalent MN | p-value No CHIP/CCUS vs CHIP/ CCUS | p-value CHIP/CCUS vs MN |
|---|---|---|---|---|---|
| Number of Cytopenias<br>0<br>1<br>2<br>3 | <br>30 (46.9%)<br>32 (50%)<br>2 (3.1%)<br>0 | <br>7 (35%)<br>13 (65 %)<br>0<br>0 | <br>0<br>1 (33.3%)<br>1 (33.3%)<br>1 (33.3%) | | |
| Anemia | 34 (53.1%) | 13 (65%) | 3 (100%) | >0.99 | 0.25 |
| Anemia + Thrombocytopenia | 2 (3.1%) | 0 | 2 (66.7%) | >0.99 | **0.01** |
| Thrombocytosis | 15 (23.4%) | 1 (0.5%) | 0 | 0.10 | >0.99 |
| Pancytopenia | 0 | 0 | 1 (33.3%) | >0.99 | 0.13 |
| CBC | N = 64 | N = 20 | N = 3 | | |
| Hemoglobin (g/dL)<br>Median [IQR] | 12.1<br>[11, 13.4] | 12.05<br>[10.85, 12.45] | **11.7**<br>**[10.9, 12.2]** | 0.31 | 0.85 |
| Mean Corpuscular Volume (fL)<br>Median [IQR] | 89.2<br>[86, 92.8] | 88.5<br>[84, 91] | 94<br>[90.1, 100.9] | 0.31 | 0.22 |
| Red Cell Distribution Width (%)<br>Median [IQR] | 13.3<br>[12.4, 14.2] | 13.6<br>[12.6, 14.8] | **15.2**<br>**[15, 16.4]** | 0.38 | 0.08 |
| Platelets (x10$^9$ cells/L)<br>Median [IQR] | 349<br>[281, 435] | 357<br>[280.5, 399.75] | **121**<br>**[105.5, 147]** | 0.66 | **0.01** |
| White Blood Cell Count (x10$^9$ cells/L)<br>Median [IQR] | 8.83<br>[7.33, 10.75] | 8.5<br>[6.83, 10.7] | 7.42<br>[4.76, 9.73] | 0.86 | 0.62 |
| Differential | n = 55 | n = 17 | n = 3 | | |
| Neutrophil % | 72<br>[66, 76.2] | 68<br>[65, 76.9] | **49**<br>**[43.3, 57.15]** | 0.63 | **0.03** |
| Neutrophil Count (x10$^9$ cells/L)<br>Median [IQR] | 6.14<br>[4.78, 8.38] | 5.63<br>[4.49, 7.56] | 2.79<br>[2.1, 4.35] | 0.51 | 0.12 |
| Lymphocyte % | 15.3<br>[11.45, 23.3] | 17<br>[12.7, 23.9] | 26<br>[23.8, 27.05] | 0.78 | 0.15 |
| Lymphocyte Count (x10$^9$ cells/L)<br>Median [IQR] | 1.5<br>[1.02, 1.94] | 1.39<br>[1.2, 1.56] | 2.09<br>[1.3, 2.61] | 0.37 | 0.36 |
| Monocyte % | 7.9<br>[6.45-11.65] | 9<br>[6.2, 11] | **23**<br>**[18.05, 27.75]** | 0.83 | **0.01** |
| Monocyte Count (x10$^9$ cells/L)<br>Median [IQR] | 0.7<br>[0.53, 1.01] | 0.67<br>[0.51, 0.71] | **2.41**<br>**[1.35, 2.59]** | 0.43 | 0.42 |
| Eosinophil % | 1.4<br>[0.7, 2.3] | 1.2<br>[0.9, 2.7] | 0<br>[0, 0] | 0.95 | 0.09 |
| Eosinophil Count (x10$^9$ cells/L)<br>Median [IQR] | 0.15<br>[0.07, 0.2] | 0.14<br>[0.1, 0.23] | 0<br>[0, 0] | 0.53 | 0.09 |
| Inflammatory markers | ESR n = 64<br>CRP n = 53 | ESR n = 19<br>CRP n = 18 | ESR n = 3<br>CRP n = 3 | | |
| Erythrocyte Sedimentation Rate (ESR) (mm/h) | 72<br>[56, 91.5] | 71<br>[49.5, 100.5] | 77<br>[42, 84] | 0.83 | 0.47 |
| C-Reactive Protein (CRP) (mg/L) | 61.1<br>[19.1, 138.93] | 57.35<br>[11.28, 127.6] | 42.1<br>[24.5, 69.95] | 0.67 | 0.53 |

Hematologic parameters are shown for the subset of individuals with CBC and inflammatory markers at disease onset prior to initiation of steroid, stratified by no CHIP/CCUS, CHIP/CCUS or prevalent MN. Cytopenia defined by WHO standards as hemoglobin <12 g/dL for women and <13 g/dL for men, platelet < 150 (x10$^9$ cells/L), and absolute neutrophil count < 1.8 (x10$^9$ cells/L). Thrombocytosis defined by WHO standard as platelet > 450 (x10$^9$ cells/L). n = 87 patients with CBC, n = 77 with differential; n per group indicated in table.

**Supplemental Table 3: Complete blood counts and cytopenia at GCA diagnosis.**

| | Total Cohort<br>n = 111 | No CHIP/CCUS<br>n = 83 | CHIP/CCUS<br>n = 28 |
|---|---|---|---|
| Number of Cytopenias | | | |
| # on TCZ or JAKi | | | |
| 0 | 67 (60.3%) | 52 (62.7%) | 15 (53.6%) |
| 1 | 38 (34.2%) | 27 (32.5%) | 11 (39.3%) |
| 2 | 4 (3.6%) | 3 (3.6%) | 1 (3.6%) |
| 3 | 2 (1.8%) | 1 (1.2%) | 1 (3.6%) |
| Anemia | 29 (26.1%) | 19 (22.9%) | 10 (35.7%) |
| # on TCZ or JAKi | 2 (1.8%) | 2 (2.4%) | 0 (0%) |
| Thrombocytopenia | 8 (7.2%) | 7 (8.4%) | 1 (3.6%) |
| # on TCZ or JAKi | 6 (5.4%) | 5 (6.0%) | 1 (3.6%) |
| Neutropenia | 1 (0.9%) | 1 (1.2%) | 0 (0%) |
| # on TCZ or JAKi | 1 (0.9%) | 1 (1.2%) | 0 (0%) |
| Anemia + Thrombocytopenia | 3 (2.7%) | 2 (2.4%) | 1 (3.6%) |
| # on TCZ or JAKi | 0 (0%) | 0 (0%) | 0 (0%) |
| Pancytopenia | 2 (1.8%) | 1 (1.2%) | 1 (3.6%) |
| # on TCZ or JAKi | 1 (0.9%) | 1 (1.2%) | 0 (0%) |
| Thrombocytosis | 0 (0%) | 0 (0%) | 0 (0%) |
| # on TCZ or JAKi | 0 (0%) | 0 (0%) | 0 (0%) |

CBC at last follow up stratified by no CHIP/CCUS or CHIP/CCUS at time of research sequencing; prevalent MN was excluded. Absolute number and proportion of individuals with cytopenias and types of cytopenias shown per group. Number of patients using tocilizumab (TCZ) (n = 23) or janus kinase inhibitor (JAKi) (n = 1) demonstrated per level of cytopenia.

**Supplemental Table 4: Cytopenias at last follow up**

| | Total Cohort<br>n = 111 | No CHIP/CCUS<br>n = 83 | CHIP/CCUS<br>n = 28 |
|---|---|---|---|
| Referral to Hematology | 22 (19.8%) | 16 (19.3%) | 6 (21.4%) |
| Reason for Referral | | | |
| Concern for malignancy | 3 (2.7%) | 2 (2.4%) | 1 (3.5%) |
| Cytopenia | 10 (9.0%) | 6 (7.2%) | 4 (14.3%) |
| Altered SFLC | 4 (3.6%) | 4 (4.8%) | 0 (0%) |
| Anti-coagulation/ VTE | 5 (4.5%) | 4 (4.8%) | 1 (3.6%) |
| Diagnostic Testing | | | |
| LN or BM biopsy | 4 (3.6%) | 1 (1.2%) | 3 (10.7%) |
| Genetic testing | 5 (4.5%) | 2 (2.4%) | 3 (10.7%) |
| Incident Clinical Hematologic Diagnoses | | | |
| ITP | 1 | 1 | 0 |
| CCUS | 1 | 1[#] | 0 |
| MDS | 0 | 0 | 0 |
| CMML | 0 | 0 | 0 |
| AML | 1 | 0 | 1*[#] |
| DLBCL | 1 | 0 | 1*[#] |
| Myeloma | 1 | 1*[#] | 0 |

Referrals to hematology, as well as reason for referral, absolute number of individuals who underwent bone marrow biopsy or genetic testing, and incident hematologic diagnoses were documented. *Indicates patient underwent lymph node or bone marrow biopsy. [#]Indicates patient underwent genetic testing.

**Supplemental Table 5: Referral to Hematology for all GCA patients.**

### Supplemental Figures

Supplemental Figure 1: Incident GCA in the UK Biobank.

Supplemental Figure 2: Vision loss in the clinical GCA cohort

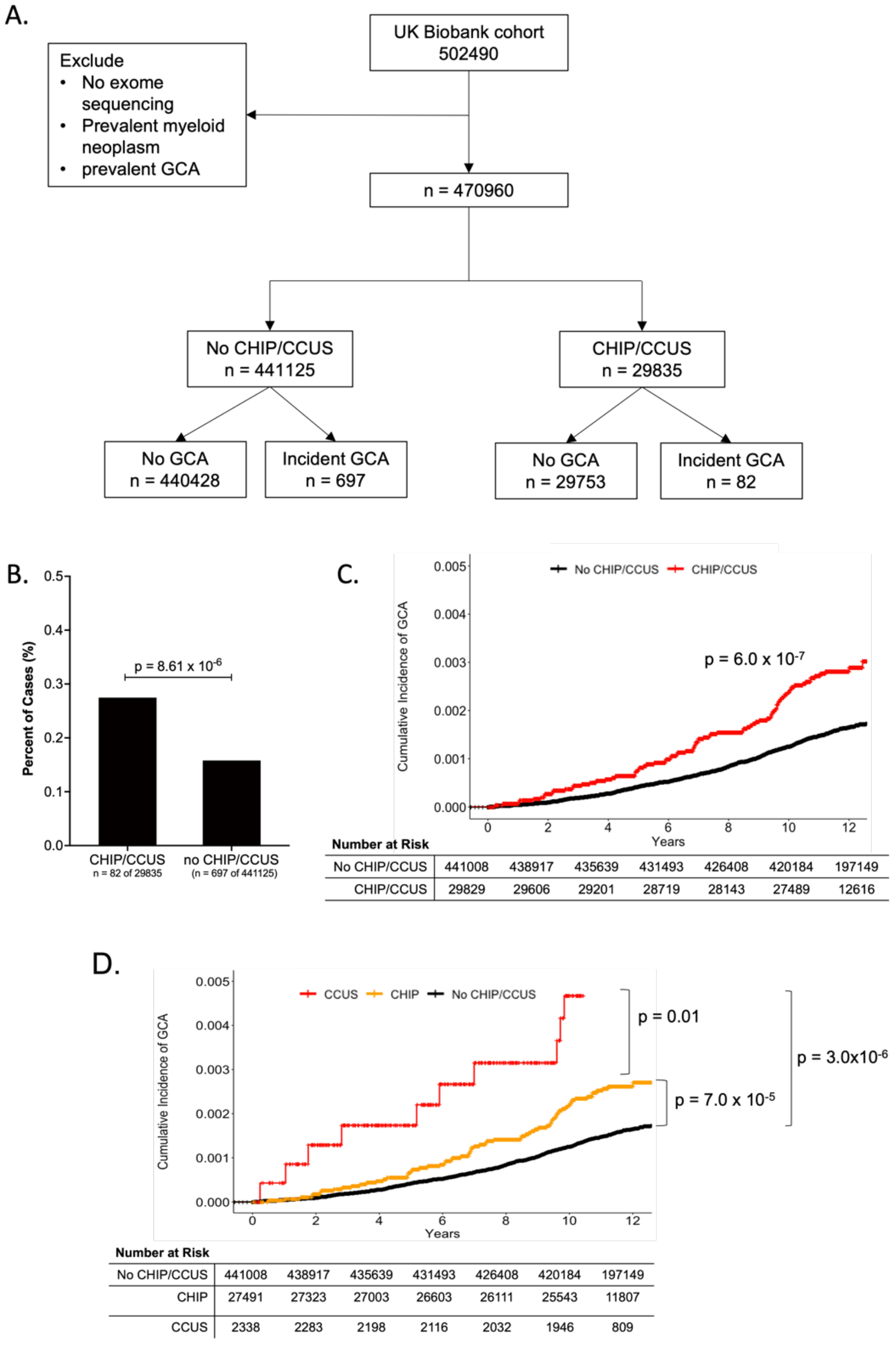


Number at Risk (C)

| | | | | | | | |
|---|---|---|---|---|---|---|---|
| No CHIP/CCUS | 441008 | 438917 | 435639 | 431493 | 426408 | 420184 | 197149 |
| CHIP/CCUS | 29829 | 29606 | 29201 | 28719 | 28143 | 27489 | 12616 |

Number at Risk (D)

| | | | | | | | |
|---|---|---|---|---|---|---|---|
| No CHIP/CCUS | 441008 | 438917 | 435639 | 431493 | 426408 | 420184 | 197149 |
| CHIP | 27491 | 27323 | 27003 | 26603 | 26111 | 25543 | 11807 |
| CCUS | 2338 | 2283 | 2198 | 2116 | 2032 | 1946 | 809 |

**Supplemental Figure 1: GCA in the UK Biobank**
**A)** Flow diagram showing identification of UKB participants with CHIP/CCUS and with incident GCA. B) proportion of participants with and without CHIP/CCUS in UKB with incident GCA. P-value determined by fisher's exact test. C) Cumulative incidence of GCA in CHIP/CCUS (red curve) vs no CHIP/CCUS (black curve). D) Cumulative incidence of GCA in individuals with CCUS, CHIP and No CHIP/CCUS. For C and D, p-values comparing curves are shown as determined by log-rank test. For all statistical tests, a p-value of < 0.05 is considered statistically significance.

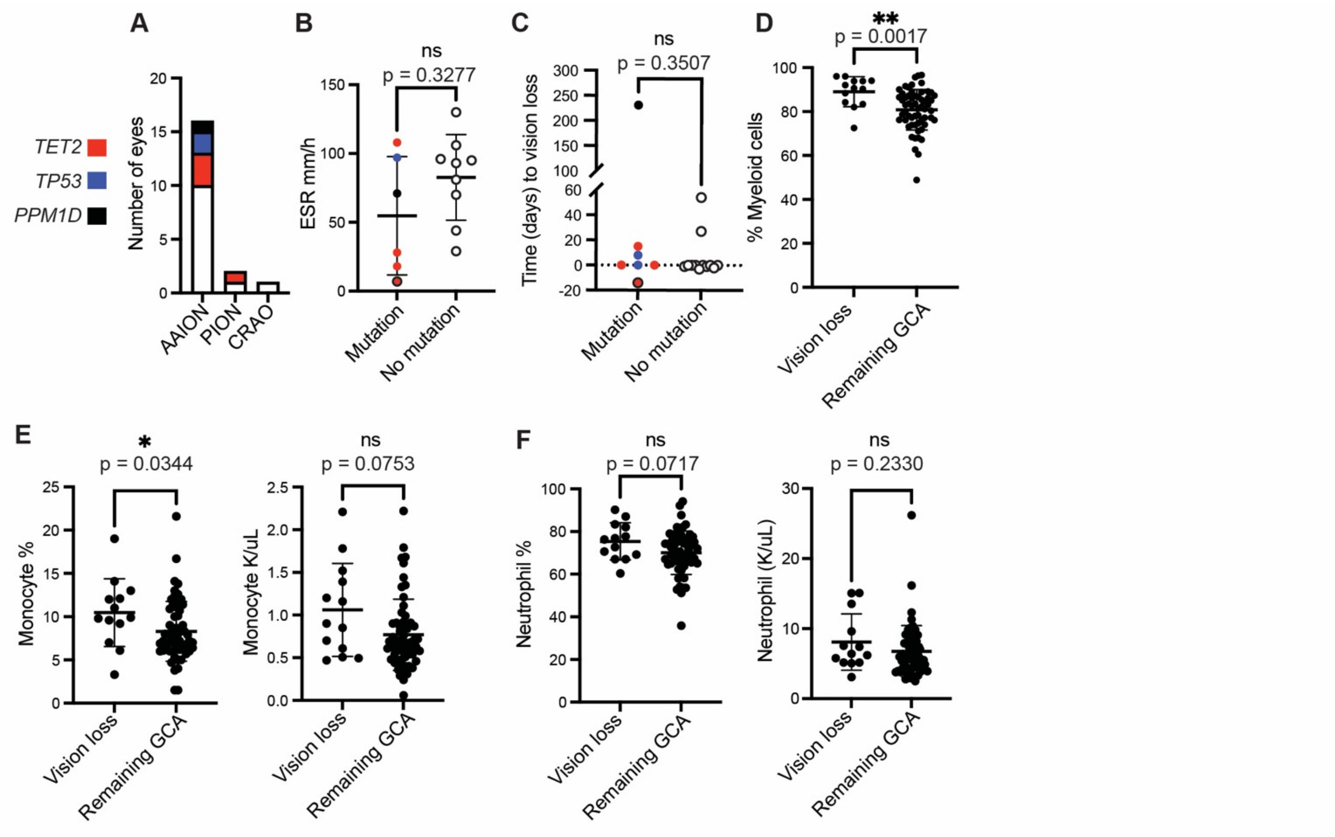


**Supplemental Figure 2: Extended analyses of vision loss in the MGB GCA cohort.**
**A)** Types of vision loss, stratified by max VAF mutation. **B)** Pre-steroid ESR for patients with vision loss. **C)** Time between vision loss and initiation of steroids for suspected GCA, where time = 0 is initiation of steroids. **C-D)** Color-code shows max VAF mutation and black outline represents individual with prevalent MN. **D-F)** Pre-steroid CBC data, excluding prevalent MN, in individuals with GCA with and without vision. **D)** Percentage myeloid cells, calculated as percentage of non-lymphoid cells. **E)** Monocyte percentage and absolute number and **F)** neutrophil percentage and absolute number.  AAION = arteritic anterior ischemic optic neuropathy. PION = posterior ischemic optic neuropathy. CRAO = central retinal artery occlusion.
